# Sex-Related Disparities in Cardiometabolic Burden Among a Large Algerian Cohort

**DOI:** 10.64898/2026.07.29.26359246

**Authors:** Mourad Boukheloua, Radia Bensemmane, Youcef Tarfani, Djamel Fourar, Houssem Baghous, Nazim Laraba, Nabil Bellik, Yehya Khlidj

**Author notes:** **Corresponding author:** Yehya Khlidj, MD, MS, mailto, Faculty of Medicine, University of Health Sciences, Algiers, Algeria. Address: Road 8 Lieutenant Mohamed Benarfa, El Biar, 16000, Algiers, Algeria.

## Abstract

**Background:** Cardiovascular diseases (CVD) are the leading cause of mortality worldwide. Historically, male sex was considered a cardiovascular risk factor however recent accumulating data suggest complex interaction between sex and CVD leading to specific inequalities affecting both males and females with specific cardiometabolic features.

**Methods:** Data from a large nationwide screening cohort were used to assess the cardiometabolic burden among males vs females in Algeria. The prevalence and adjusted Odds Ratio (aOR) of major CVD diseases were compared across the study cohorts. Records of systolic (SBP) and diastolic blood pressure (DBP), as well as findings of clinical examination, ECG, and echocardiography were collected and reviewed for analysis.

**Results:** A total of 21,522 patients were included among them 12,015 were females and 9,507 were males. The latter group had higher age (median age: 60 [51.00-68.00] vs 57 [49.00-65.00] years; p< 0.001), sitting DBP (83 [76.00-92.00] vs 80 [75.00-90.00]; p< 0.001), standing SBP (142 [130.00-160.00] vs 140 [124.00-154.00] mmHg; p< 0.001), smoking (22.1% vs 0.9%; p<0.001) and unknown diabetes (15.2% vs 13.2%; p< 0.001) rates. Moreover, females showed an overall worse cardiometabolic profile while males had a greater burden of ischemic heart disease, arrhythmia, conduction abnormalities, and left ventricular dysfunction. On multivariate analysis, female sex was correlated with higher aOR for known hypertension (aOR:1.55; 95%CI:1.40-1.72; p<0.001) and known dyslipidaemia (aOR: 1.23; 95%CI:1.09-1.39; p=0.001). On the other hand, female sex predicted lower risk of any CVD (aOR:0.84; 95%CI:0.72-0.98; p=0.028) and known cardiovascular disease (aOR: 0.79; 95%CI: 0.67-0.94; p=0.007).

**Conclusion:** CVD affect males and females differently which reflects multifaceted biological, cardiometabolic, and sociodemographic disparities. These findings highlight the need to reduce the sex gap in the prevention and management of CVD.

## 1. Introduction

Cardiovascular disease (CVD) remains the leading cause of death worldwide. According to the latest estimates from the Global Burden of Disease, the median age-standardised death rate per 100 000 population of CVD reached 215.2 (194.3 to 229.8) in 2023 compared to 322.0 (298.9 to 340.4) in 2000, thus accounting for 33.2% (37.9 to 28.3) decrease in attributed mortality (1). The adherence rates to guideline recommendations have also improved over time which directly mitigated cardiovascular events (2). This highlights the effectiveness of prevention and management strategies implemented in the last two decades in lowering the burden of CVD. However, more efforts are still largely needed to promote cardiovascular health, particularly given that decreasing cardiovascular mortality has decelerated and is not meeting the warranted level. Factors that cause this are suboptimal control of cardiovascular risk factors notably hyperglycemia, increased blood pressure, obesity, alcohol consumption, and dyslipidemia, alongside persistent socioeconomic disparities (3). In line with this, reducing inequalities across various sociodemographic subgroups appears as one of the fundamental strategies to lower the burden of CVD and enhance tailored care for more vulnerable individuals.

Notably, recent research has focused on the sex gap in CVD and the underrepresentation of females in cardiovascular care and trials (4,5). Indeed, the burden of ischemic heart disease, hypertension, obesity, and diabetes was shown to differ by sex with possibly higher prevalence in males but greater morbimortality in females (6,7). The interventional outcomes of cardiovascular procedures also seem to be less optimal among females patients compared to males for similar comorbidity status (5). On the other hand, other studies showed male sex to be associated with worse CVD risk profile and greater rates of all-cause and cardiovascular mortality in both the general and cardiovascular population with CVD (8,9).

Importantly, this evidence comes predominantly from Western data and can not be generalized in the absence of multinational investigations. Oppositely, recent studies from African populations revealed minimal sex-based disparities in cardiovascular outcomes (10). In this background, the status of the sex gap in CVD among underrepresented populations such as those of North Africa remains scarce and requires special investigations. Therefore, we conducted the present large-scale population-level analysis.

## 2. Methods

### 2.1. Study design, database, and participants

We used data from a large nationwide screening cohort to assess the cardiometabolic burden among males vs females in Algeria. During the period of 2015-2025, the Algerian Ministry of Health led a national program for Non-communicable Disease Prevention Path to evaluate the current burden of non-communicable diseases among the general population through an extensive screening campaign. Participants were informed through media platforms and were recruited randomly to achieve maximum real-world evaluation of the cardiovascular and metabolic status locally.

Cardiometabolic diagnoses and outcomes were based on patient claims, clinical examination, laboratory workup, ECG, and echocardiography, all performed firsthand by a multidisciplinary team comprising cardiologists, general practitioners, and laboratory specialists, who were transported throughout the country regions by highly equipped mobile clinics.

The included participants were any adult individuals aged ≥18 years who agreed to benefit from free medical visits by the multidisciplinary team. These participants were from all regions across the nation, both urban and rural. Before the examination, the participants medical history was not known by the examiners to avoid selective recruitment.

### 2.2. Statistical analysis

Continuous variables were summarized as median (interquartile range [IQR]), while categorical variables were presented as counts and percentages. Because several continuous measures exhibited non-normal distributions, between-sex comparisons for continuous variables were conducted using the Mann–Whitney U test. Comparisons of categorical variables between men and women were performed using the χ² test. All tests were two-sided, and a p value < 0.05 was considered statistically significant.

Importantly, due to factors such as incomplete data reporting or recording (eg, uncertainty of some participants about their medical information), and the inability to complete the full screening protocol by some participants, the final data were missing for certain variables. Because of incomplete reporting or incomplete screening procedures, missingness varied by variable. The sample size (N) used for each analytic model is reported in the **Supplementary material.**

To evaluate whether sex was independently associated with major cardiometabolic outcomes, we fitted multivariable regression models. For binary outcomes, binomial logistic regression was used and results were reported as adjusted odds ratios (aORs) with 95% confidence intervals (CIs). Age and major cardiometabolic covariates were included based on clinical relevance and outcome-specific considerations (e.g., age, body mass index[BMI], and key comorbidities such as diabetes, hypertension, dyslipidemia, and family history when applicable). For continuous outcomes (e.g., sitting systolic blood pressure[SBP]), multivariable linear regression was applied, and results were reported as adjusted β coefficients (with corresponding 95% CIs). Logistic model fit was summarized using deviance, Akaike information criterion (AIC), and McFadden’s pseudo-R². Cumulative prevalence for major cardiovascular and metabolic conditions was estimated according to age and compared between sexes using Gray’s test. All analyses were performed using R software (version 4.5.2).

### 2.3. Ethical approval

This study was conducted under the supervision of the Algerian Ministry of Health, and with the approval of the National Committee for Health Ethics. Informed and written consent was obtained from all study participants. The investigations were conducted in accordance with the principles outlined in the Declaration of Helsinki (1975, revised in 2013).

## 3. Results

### 3.1. Sociodemographic and clinical characteristics

A total of 21,522 adults were included (12,015 females and 9,507 males). Males were significantly older than females (median age 60 [51–68] vs 57 [49–65] years; p < 0.001). Compared with males, females had greater adiposity, with higher BMI (median 29.30 [25.78–32.99] vs 26.56 [23.84–29.41] kg/m²; p < 0.001), while waist circumference differed modestly (median 100 [92–110] vs 100 [91–107] cm; p < 0.001). Hemodynamic parameters also differed thus males demonstrated higher blood pressure values (e.g., median sitting SBP: 140 [130–160] vs 140 [121–153] mmHg; p < 0.001, and median sitting diastolic blood pressure [DBP]: 83 [76–92] vs 80 [75–90] mmHg; p < 0.001), whereas females had higher median sitting heart rate (80 [72–88] vs 78 [69–86] bpm; p < 0.001). Glycemic indices were comparable between sexes (median HbA1c level: 7.0 [6.0–9.0] in both; p = 0.573; median capillary blood glucose level: 1.45 [1.10–2.38] vs 1.50 [1.10–2.43]; p = 0.078), while creatinine levels were higher in males (median: 8.0 [6.0–11.0] vs 7.0 [5.0–10.0]; p < 0.001). Lipid measures (LDL, HDL, total cholesterol, and triglycerides) did not differ significantly (**Table 1**).

**Table 1.** Baseline demographics and clinical parameters by sex.

| Variable | Females | Males | P value |
| --- | --- | --- | --- |
| Age, years | 57 [49.00–65.00],<br>n=12015 | 60 [51.00–68.00],<br>n=9507 | <.001 |
| BMI, kg/m<sup>2</sup> | 29.30 [25.78–<br>32.99], n=8252 | 26.56 [23.84–<br>29.41], n=6655 | <.001 |
| Waist<br>circumference, cm | 100.00 [92.00–<br>110.00], n=8252 | 100.00 [91.00–<br>107.00], n=6655 | <.001 |
| Sitting SBP, mmHg | 140 [121.00–<br>153.00], n=3687 | 140 [130.00–<br>160.00], n=3035 | <.001 |
| Sitting DBP, mmHg | 80 [75.00–90.00],<br>n=3687 | 83 [76.00–92.00],<br>n=3035 | <.001 |
| Sitting heart rate,<br>bpm | 80 [72.00–88.00],<br>n=3687 | 78 [69.00–86.00],<br>n=3035 | <.001 |
| Standing SBP,<br>mmHg | 140 [124.00–<br>154.00], n=3687 | 142 [130.00–<br>160.00], n=3035 | <.001 |
| HbA1c | 7.00 [6.00–9.00],<br>n=633 | 7.00 [6.00–9.00],<br>n=598 | 0.573 |
| Capillary blood<br>glucose | 1.45 [1.10–2.38],<br>n=11546 | 1.50 [1.10–2.43],<br>n=9098 | 0.078 |
| Microalbuminuria | 5.00 [5.00–56.00],<br>n=734 | 5.00 [5.00–87.00],<br>n=627 | 0.092 |
| Creatinine | 7.00 [5.00–10.00],<br>n=507 | 8.00 [6.00–11.00],<br>n=488 | <.001 |
| LDL | 1.00 [1.00–1.00],<br>n=124 | 1.00 [1.00–1.00],<br>n=123 | 0.200 |
| HDL | 1.00 [1.00–1.00],<br>n=53 | 1.00 [1.00–1.00],<br>n=70 | 0.612 |
| Total cholesterol | 1.00 [1.00–2.00],<br>n=28 | 1.00 [1.00–1.25],<br>n=40 | 0.369 |
| Triglycerides | 1.00 [1.00–1.00],<br>n=81 | 1.00 [1.00–2.25],<br>n=40 | 0.355 |

Values are median [25th–75th percentile], n as available per variable. P values from Mann–Whitney U tests.

### 3.2. Sex Differences in Cumulative Prevalence of CVD

Age-specific cumulative prevalence curves for total cardiovascular disease, coronary heart disease, stroke, and heart failure are presented per sex in **Figure 1** (n=9,507 males and 12,015 females). Gray’s test p values are shown in the corresponding panels. The cumulative prevalence for total CVD, coronary artery disease, stroke, hypertension, diabetes, and peripheral artery disease showed a sharp age-dependent increase, with the curves markedly accelerating after ∼40–50 years. Subsequently, a milder acceleration in the cumulative prevalence was noted for heart failure, showing a more progressive increase with a delayed surge until 70 years. In terms of sex, there was a statistically significant earlier onset of coronary artery disease in males at 37–40 years (P < 0.001). However, this sex bias becomes less distinguishable at higher ages. Peripheral artery disease also demonstrated an earlier onset at the age of 35 however, with increased age, the male predominance became inconsistent, with females having higher cumulative prevalence at the age of 45-55 then males displaying the highest rates afterwards (P=0.025). The occurrence of hypertension, dyslipidemia, and diabetes was prematurely observed in females at around the ages of 40, 50, and 45 years, respectively (P < 0.001 for all), with the sex gap expanding over age. No significant differences were observed for the age-specific cumulative prevalence of CVD (P = 0.149), stroke (P = 0.359), and heart failure (P = 0.533) across sex groups.

**Figure 1.**
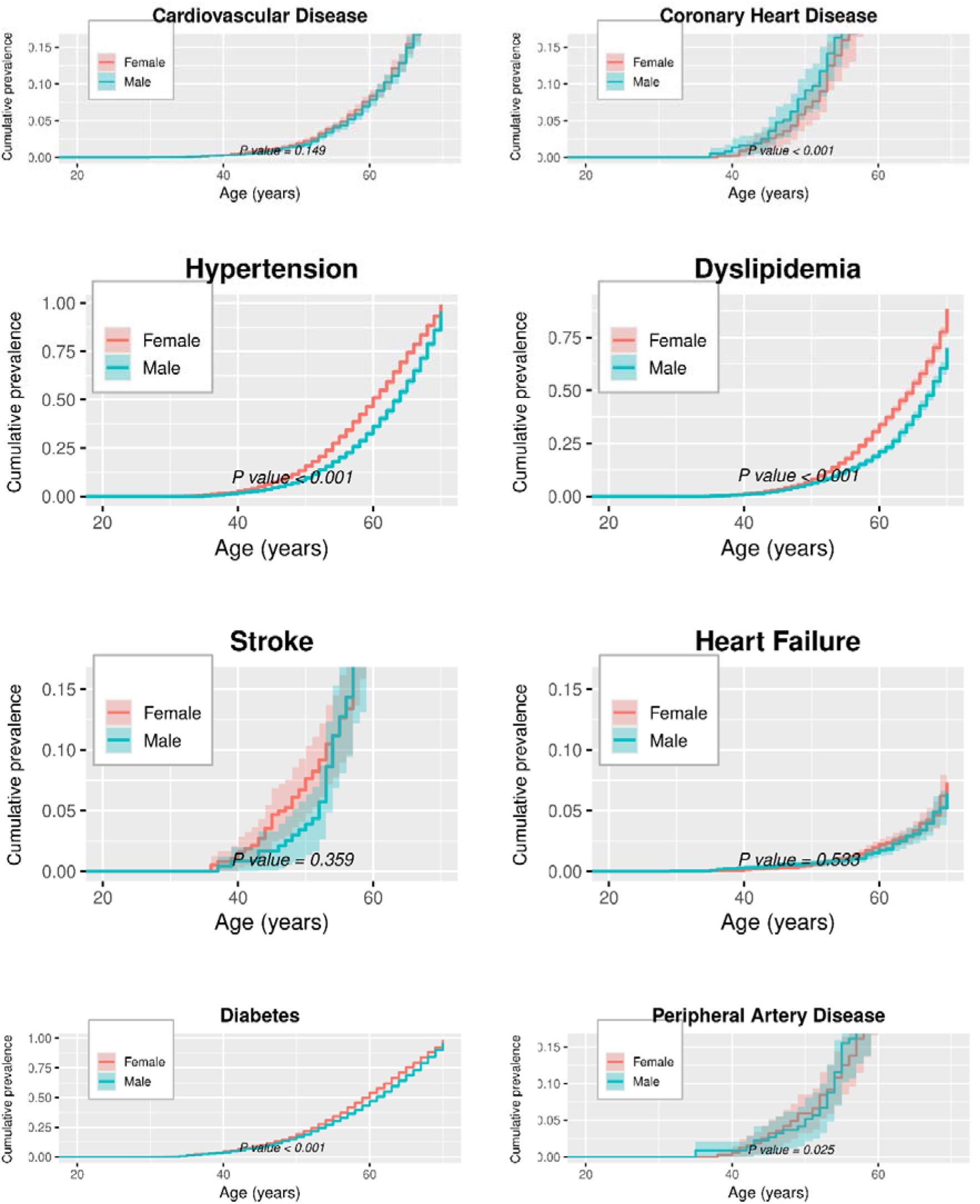
Descriptive age-specific cumulative prevalence analysis for total cardiovascular disease, coronary heart disease, stroke, and heart failure by sex. Panels display cumulative prevalence stratified by sex (n=9,507 males and 12,015 females). P values obtained from Gray’s test.

### 3.3. Cardiometabolic risk factors and prevalent disease burden

Females more frequently reported several established cardiometabolic conditions. Known hypertension was more common among females (72.0%) than males (61.5%; p < 0.001), and so was known dyslipidemia (38.7% vs 29.5%; p < 0.001) and known diabetes (70.1% vs 68.5%; p = 0.030). Consistent with anthropometric differences, abdominal obesity was more common in females (22.9%) compared with males (14.2%; p < 0.001). In contrast, current smoking showed a marked male predominance (22.1% in males vs 0.9% in females; p < 0.001). Remarkably, family history of hypertension (59.4% vs 48.4%; p<0 .001), diabetes (57.8% vs 49.9%; p<0.001), and dyslipidemia (30.8% vs 21.0%; p<0.001) were more frequently reported in the female cohort. Importantly, in both sexes there was a considerable proportion of unknown diabetes and diabetic foot, with slightly higher rates in males (15.2% vs 13.2%; p<0.001 and 26.6% vs 24.0%; p=0.022, respectively).

Markers of renal involvement showed modest sex differences, notably proteinuria (9.6% for males vs 8.3% for females; p = 0.013) and microalbuminuria (12.7% for males vs 10.8%; p = 0.001 for females) were more frequent in males, while chronic kidney disease prevalence was similar between sexes (2.0% for males vs 1.9% for females; p = 0.540) (**Table 2**).

**Table 2.** Cardiometabolic risk factors and comorbidities by sex.

| Variable | Females | Males | P value |
| --- | --- | --- | --- |
| known Diabetes | 5631/8029 (70.1%) | 4416/6450 (68.5%) | 0.030 |
| Known hypertension | 5814/8072 (72.0%) | 3665/5958 (61.5%) | $< 0.001$ |
| Known Dyslipidemia | 2325/6001 (38.7%) | 1372/4649 (29.5%) | $< 0.001$ |
| Abdominal obesity | 1067/4662 (22.9%) | 554/3889 (14.2%) | $< 0.001$ |
| Smoking | 84/9661 (0.9%) | 1715/7750 (22.1%) | $< 0.001$ |
| Hypertension risk factors | 5247/10007 (52.4%) | 3278/7709 (42.5%) | $< 0.001$ |
| Risk factors of diabetes | 5123/10038 (51.0%) | 3868/7778 (49.7%) | 0.084 |
| Dyslipidemia risk factors | 2667/9529 (28.0%) | 1682/7284 (23.1%) | $< 0.001$ |
| Family history of hypertension | 4005/6742 (59.4%) | 2579/5327 (48.4%) | <□.001 |
| Family history of diabetes | 3890/6735 (57.8%) | 2661/5337 (49.9%) | <□.001 |
| Family history of dyslipidemia | 2037/6603 (30.8%) | 1099/5236 (21.0%) | <□.001 |
| Unknown diabetes | 1212/9167 (13.2%) | 1084/7136 (15.2%) | <□.001 |
| chronic kidney disease | 142/7619 (1.9%) | 118/5870 (2.0%) | 0.540 |
| Proteinuria | 570/6886 (8.3%) | 508/5315 (9.6%) | 0.013 |
| Micro-albuminuria | 742/6891 (10.8%) | 668/5280 (12.7%) | 0.001 |
| Diabetic foot | 770/3214 (24.0%) | 711/2677 (26.6%) | 0.022 |

Values are n/N (%). P values from $\chi^2$ tests.

Furthermore, the rates of known cardiovascular disease were slightly higher in males (10.4% vs 9.2%; p=0.011), with this group showing a much more pronounced predominance in terms of coronary artery disease (65.8% vs 46.7%; p<0.001) and peripheral artery disease (35.3% vs 27.8%; p=0.029). Comparable rates of stroke history were reported by females and males (29.7% vs 35.7%; p=0.068). Regarding symptoms and clinical burden, females more frequently reported dyspnea (17.4% vs 13.7%; p < 0.001), whereas angina did not differ significantly (p = 0.152) (**Table 3**).

**Table 3.**
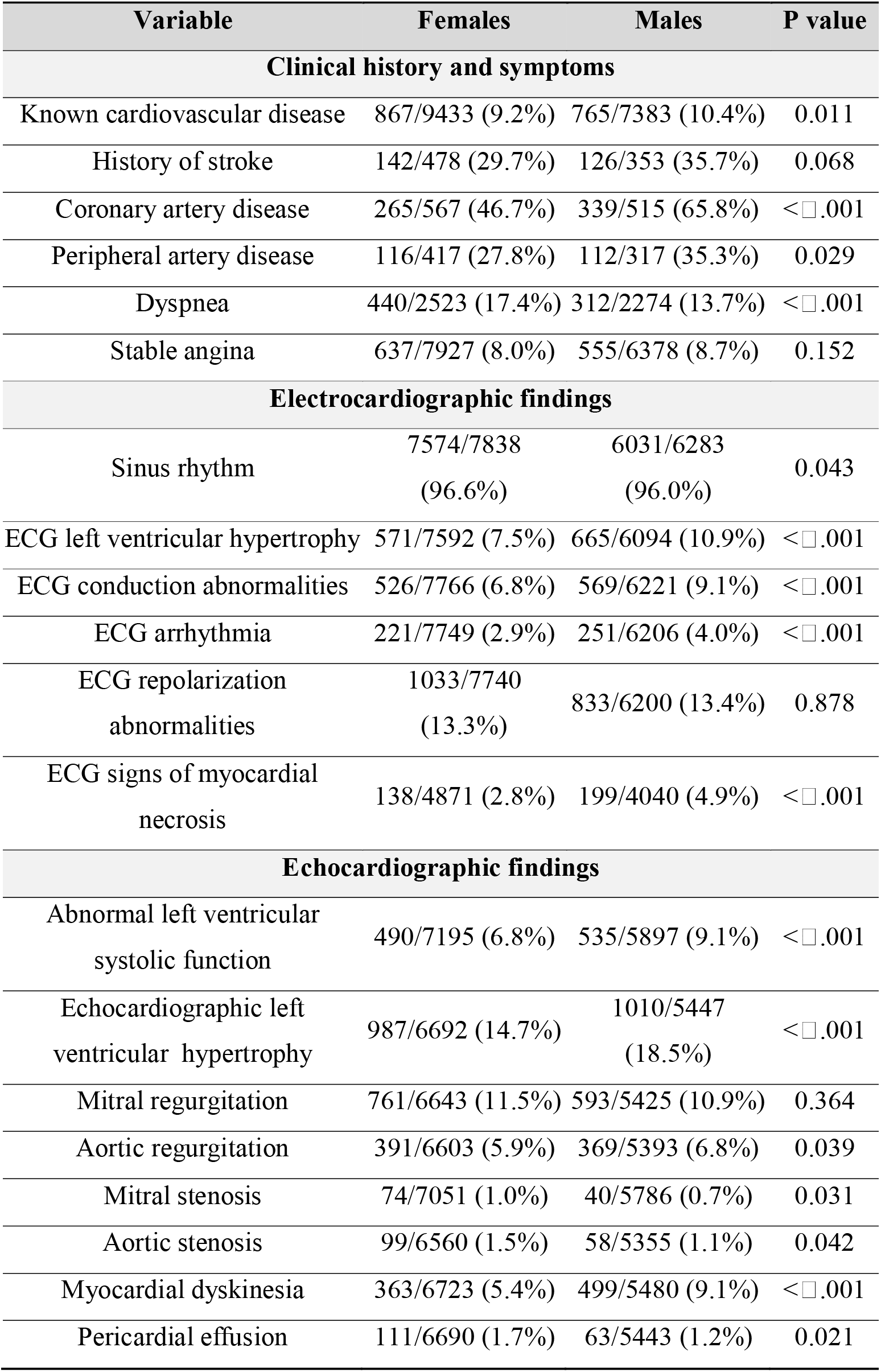
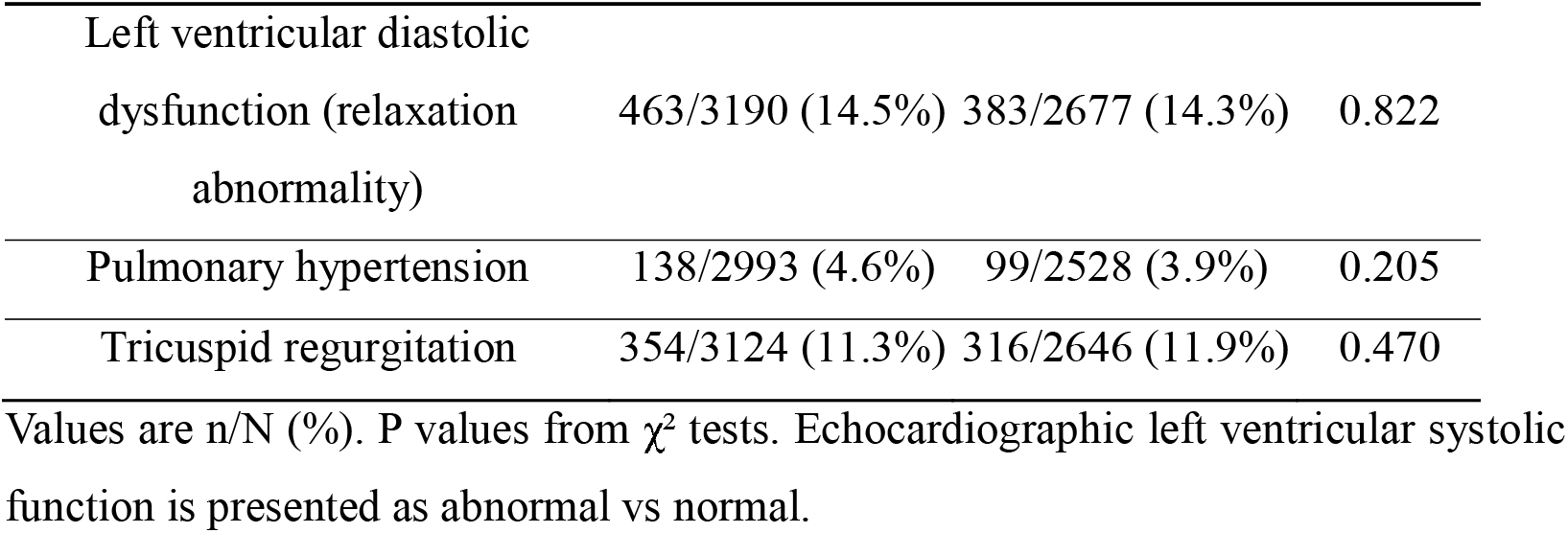
Cardiovascular history, ECG, and echocardiographic findings by sex.

### 3.4. ECG and echocardiographic profile

Across electrophysiological and echocardiographic assessments, males had a higher prevalence of several abnormalities (**Table 3**). On ECG, males more often demonstrated left ventricular hypertrophy (10.9% vs 7.5%; p < 0.001), conduction abnormalities (9.1% vs 6.8%; p < 0.001), arrhythmia (4.0% vs 2.9%; p < 0.001) and signs of myocardial necrosis (4.9% vs 2.8%; p<0.001). Echocardiographic left ventricular hypertrophy was also more frequent in males than in females (18.5% vs 14.7%; p < 0.001), as were abnormal left ventricular systolic function (9.1% vs 6.8%; p < 0.001), aortic regurgitation (6.8% vs 5.9%; p=0.039), and myocardial dyskinesia (9.1% vs 5.4%; p<0.001). Minimal but significant differences were observed in females vs males regarding the rates of mitral stenosis (1.0% vs 0.7%; p=0.031), aortic stenosis (1.5% vs 1.1%; p=0.042), and pericardial effusion (1.7% vs 1.2%; p=0.021). Pulmonary hypertension did not differ by sex (p = 0.205).

### 3.5. Patterns of cardiovascular risk management

Sex differences were also observed in the clinical management of CVD. Among participants with chest pain, female sex was associated with a higher rate of unexplored symptoms (62.5%) and a lower rate of explored symptoms (19.0%) than male sex (51.5% and 30.7%, respectively; overall p = 0.026). The proportion managed by a cardiologist was similar between females and males (71.9% vs 72.2%; p = 0.775). Among patients with treated hypertension, intensification patterns (mono-, dual-, or triple therapy) were comparable across sex groups (overall p = 0.281), although the distribution of antihypertensive drug classes differed modestly. Hence, the prescription rates of angiotensin II receptor blockers (ARBs) (56.8% vs 60.2%), angiotensin-converting enzyme (ACE) inhibitors (13.1% vs 14.4%), calcium channel blockers (10.9% vs 11.8%), and diuretic therapy (2.8% vs 3.0%) were slightly lower in females (overall p = 0.017). Diabetes treatment also differed across sex groups, with oral antidiabetic drugs prescribed more frequently in males (70.6% vs 65.4%), whereas combination oral therapy plus insulin was more common in females (18.3% vs 14.6%; overall p=0.014). Lipid-lowering therapy among patients with dyslipidemia did not significantly differ by sex (p = 0.413) (**Table 4**).

**Table 4.**
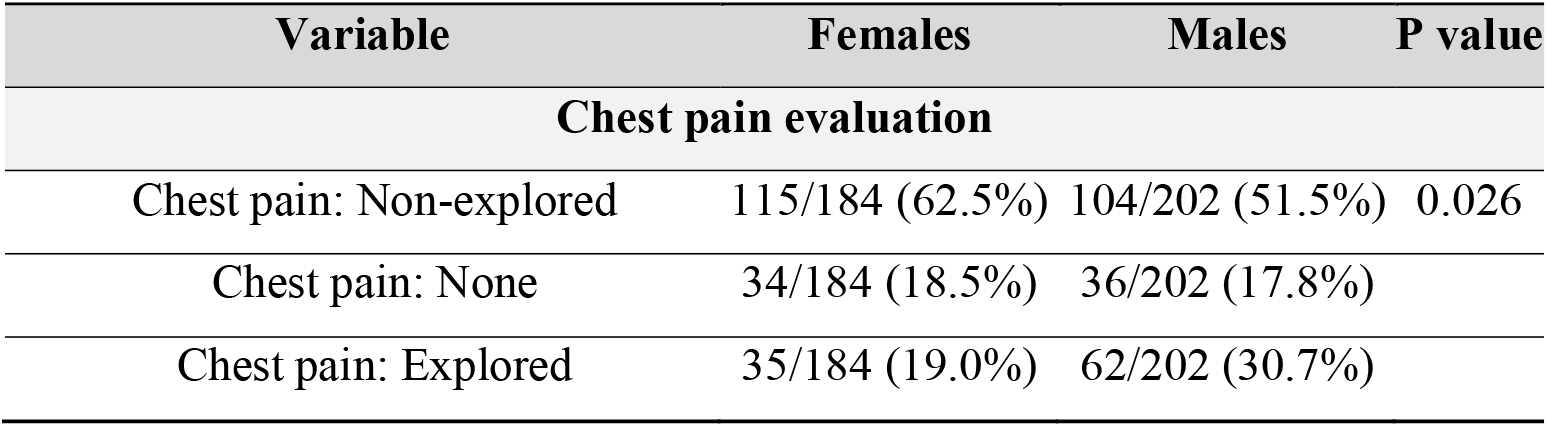
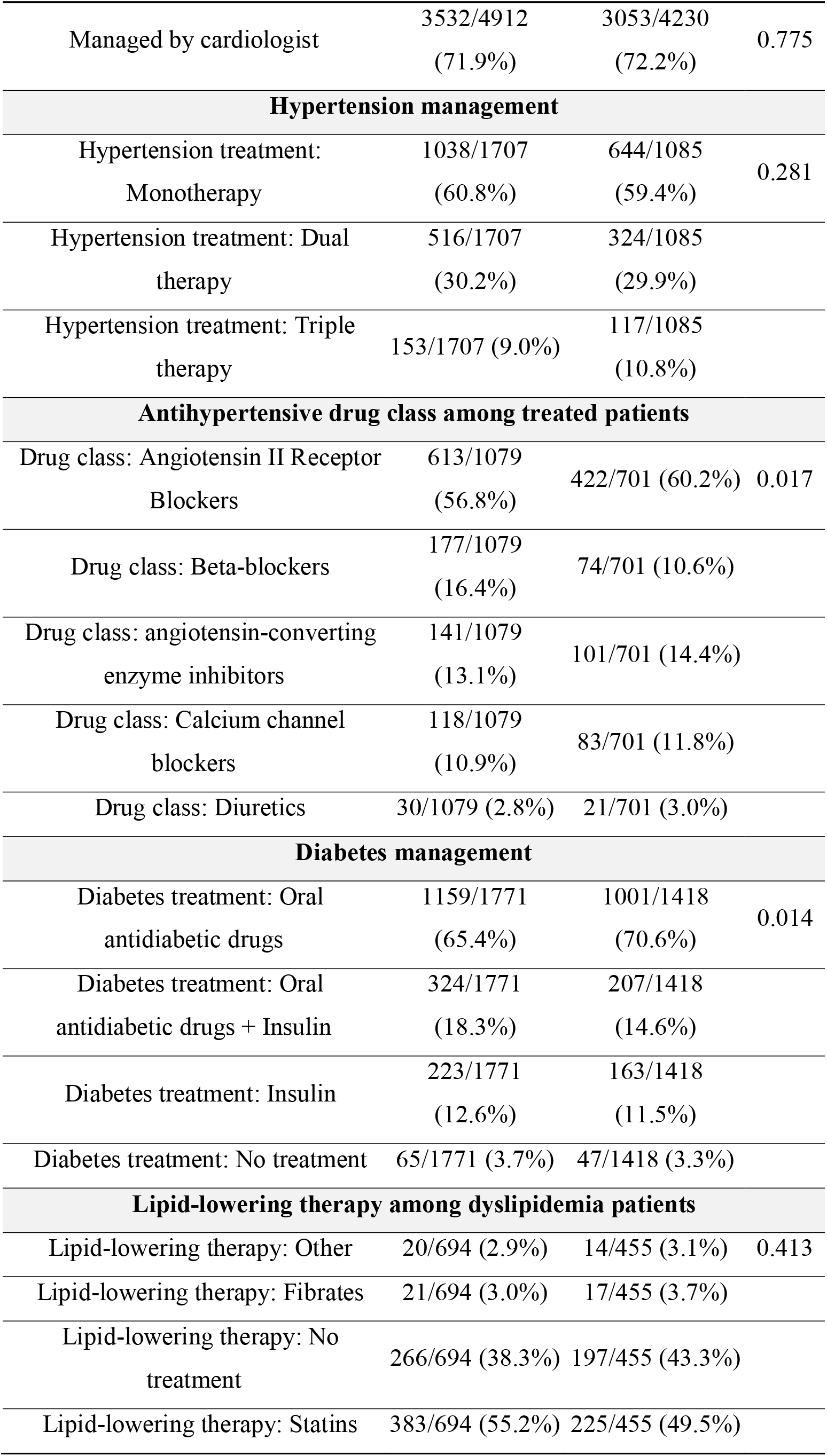
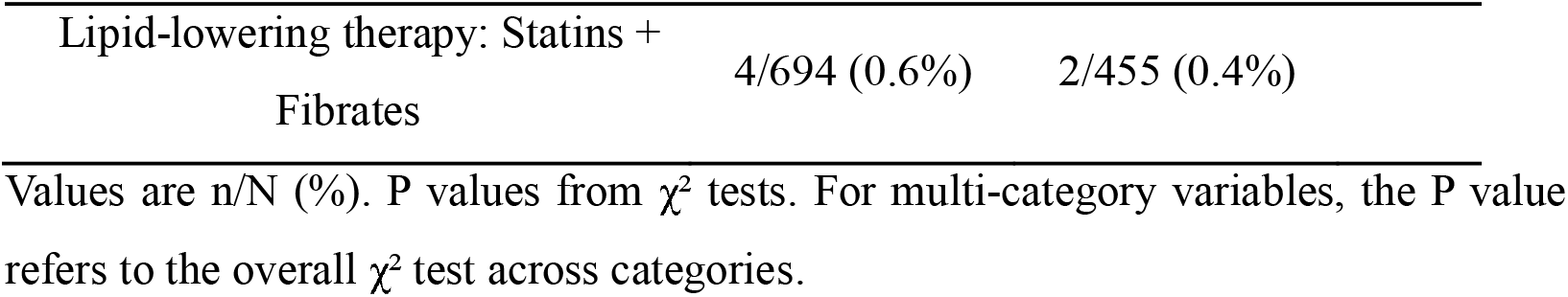
Patterns of cardiovascular risk management by sex.

### 3.6. Multivariable regression analysis

In multivariable logistic regression models adjusted for age and clinically relevant cardiometabolic covariates, female sex was independently associated with higher odds of known hypertension (aOR 1.55, 95% CI 1.40–1.72; p < 0.001) and known dyslipidemia (aOR 1.23, 95% CI 1.09–1.39; p = 0.001), and with lower odds of any CVD (aOR 0.84, 95% CI 0.72–0.98; p = 0.028) and known cardiovascular disease (aOR 0.79, 95% CI 0.67–0.94; p = 0.007). Sex was not independently associated with diabetes (aOR 0.97, 95% CI 0.87–1.08; p = 0.584) and chronic kidney disease (aOR 1.16, 95% CI 0.79–1.69; p = 0.458) (**Table 5**; **Figure 2**). For continuous hemodynamic measures, males had a higher age- and BMI-adjusted sitting SBP (β = +4.30 mmHg; 95% CI 3.19 to 5.41; p < 0.001) in linear regression (Supplementary material).

**Figure 2.**
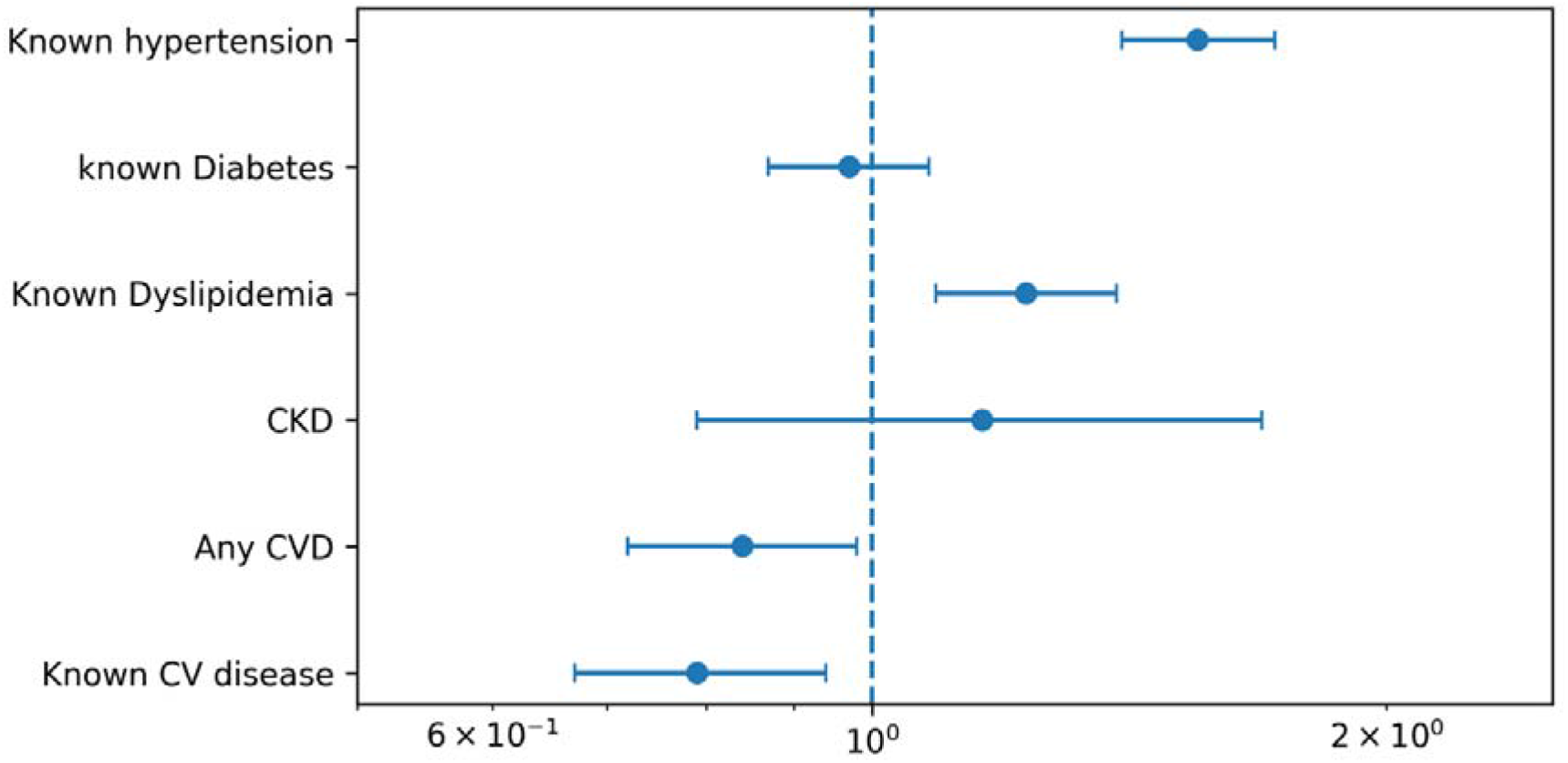
Forest plot of adjusted odds ratios for females vs males across selected cardiovascular outcomes. Adjusted odds ratios and 95% CIs correspond to Table 5. The vertical dashed line indicates aOR=1.

**Figure 3.**
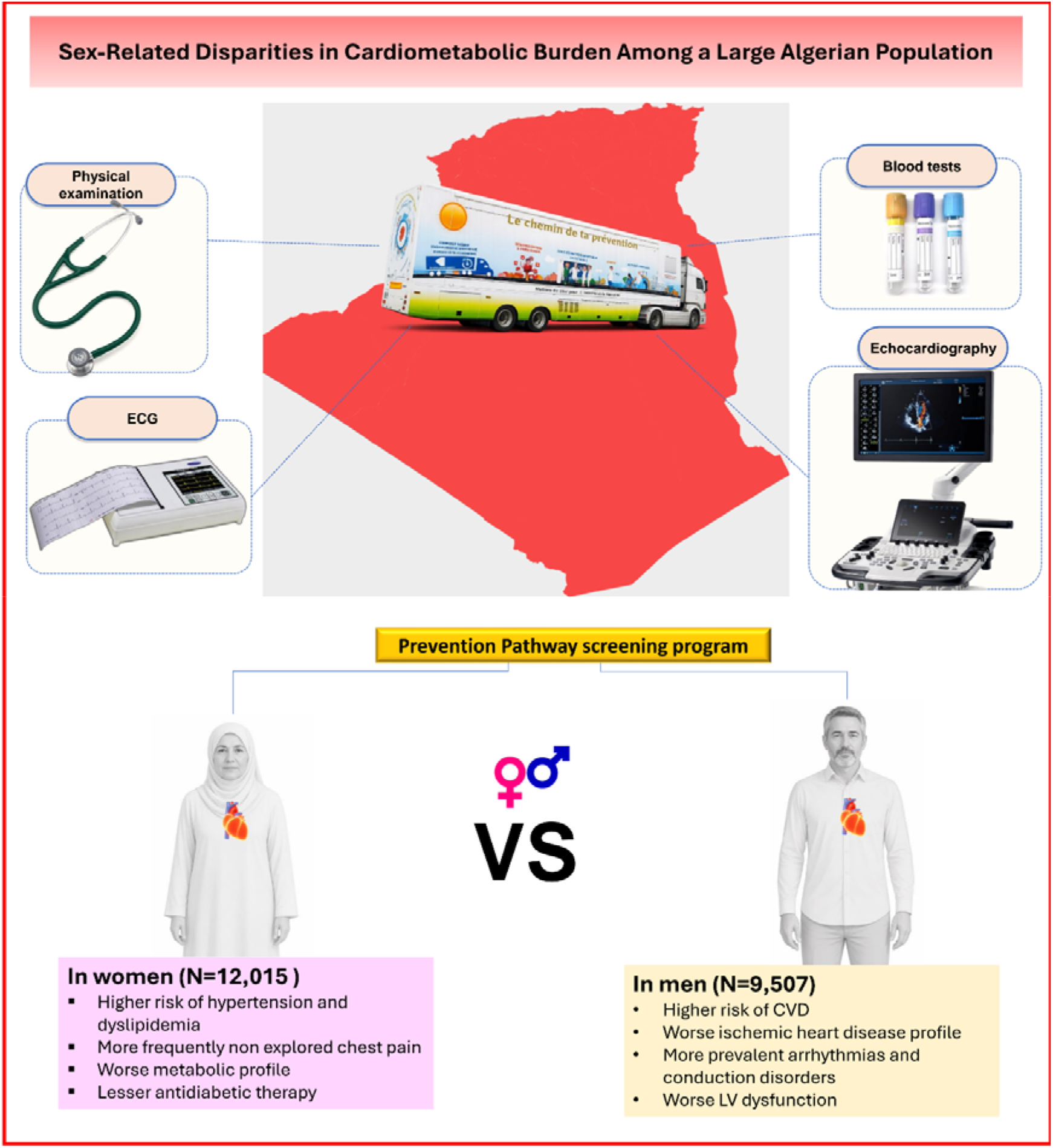
Graphical Abstract.

**Table 5.** Multivariable logistic regression: association of female sex with selected outcomes.

| Outcome | aOR<br>(Females vs<br>Males) | 95% CI | P value | Covariates in the model |
| --- | --- | --- | --- | --- |
| Known hypertension | 1.55 | 1.40–1.72 | <.001 | Age, BMI, known Diabetes, Known Dyslipidemia |
| known Diabetes | 0.97 | 0.87–1.08 | 0.584 | Known Dyslipidemia, Family history of diabetes, BMI |
| Known Dyslipidemia | 1.23 | 1.09–1.39 | 0.001 | known Diabetes, known hypertension |
| Chronic kidney disease | 1.16 | 0.79–1.69 | 0.458 | Known hypertension, known Diabetes, Known Dyslipidemia |
| Any CVD | 0.84 | 0.72–0.98 | 0.028 | Age, BMI, Known hypertension, Known<br>Dyslipidemia |
| Known<br>cardiovascular<br>disease | 0.79 | 0.67–0.94 | 0.007 | Known hypertension, Known<br>Dyslipidemia |

aOR indicates adjusted odds ratio for females vs males. Covariates are as specified in each model output.

## 4. Discussion

### 4.1. Summary of the findings

In this large study based on records of a nationwide screening campaign, sex was a critical determinant of cardiovascular and metabolic burden, with males and females showing various distinctive features suggesting the sex gap notion in North African populations. Notably, the female group displayed a greater prevalence of hypertension, dyslipidemia, diabetes, and abdominal obesity, suggesting a greater metabolic burden. In contrast, male sex was correlated with higher rates of coronary and peripheral artery disease, left ventricular structural and functional anomalies, arrhythmias, and conduction disorders, likely reflecting a more ischemic phenotype. On multivariate analysis, this distinction persisted, with females demonstrating an increased likelihood of hypertension and known dyslipidemia but a lower risk of overall CVD, which is likely due to the lower prevalence of coronary artery disease and more delayed disease onset as observed in the age-specific cumulative prevalence subanalysis (**graphical abstract**).

### 4.2. Worse metabolic and anthropometric status in females

Females showed additional risk and earlier onset of hypertension and dyslipidemia alongside a more obese phenotype, as assessed by BMI and waist circumference, both being increased in this group compared to male counterparts. Additionally, female sex was associated with a younger age of diabetes as well as a higher prevalence of diagnosed cases, although the latter correlation has disappeared after adjustment for known dyslipidemia, family history of diabetes, and BMI. According to Western data, females have a greater quantitative and qualitative deficiency in cardioprotective physical activity which would favor overweight and excessive fat accumulation compared to males (11). In the Middle East and North Africa (MENA) region, this inequality is also present, perhaps to a greater extent, echoing numerous sociocultural barriers that may favor sedentary behavior among females (12). The elevated prevalence of hypertension in females observed in our study is in accordance with previous MENA studies (13,14). Nevertheless, the sex-based difference in hypertension appears to follow age-specific epidemiology: at age 40, females are more affected than males, whereas before this age, the opposite is observed (13). Oppositely, Western data suggest a net male predominance for hypertension (15). Such heterogeneity would result from multiple sociodemographic and ethno-geographic differences but also from different diagnostic strategies and criteria.

Among females, the dyslipidemic status and epidemiology are shaped by multifaceted factors such as hormonal phase (eg, pregnancy and menopause), pathophysiological phenotype (primary or secondary), sociocultural and healthcare bias (underrepresentation and undertreatment) (16). One recent meta-analysis of 206 studies found a higher prevalence of hypertriglyceridemia among males, and inversely, greater rates of low HDL and high LDL dyslipidemia among females. Another relevant finding was that Algeria had among the highest prevalences of female-dominated dyslipidemia (ie, low HDL) as well as hypercholesterolemia, compared to other global rates, suggesting a persistent burden of dyslipidemia with the need for more equitable interventions targeting Algerian females (17).

### 4.3. Earlier onset and higher burden of coronary artery disease in males

Males in our study had a more accelerated coronary artery disease onset, starting to be noticed at around 37 years of age, and persisted in the following age groups. This finding agrees with the recent observation of Freedman et al., who reported a 7-year difference between males and females in terms of the age at which a 5% incidence of CVD is reached (4). Lifestyle habits could represent a major player in this difference, with male-associated behaviors linked to more precipitating risk factors such as smoking and proatherogenic dietary features (18). However, such factors alone do not explain the specific higher tendency to develop coronary artery disease in males. Thus, biological characteristics in males determine this risk through multiple mechanisms, including missed estrogenic inhibitory effects against coronary artery disease and atherogenesis (19), Y chromosome-related consequences on coronary health (20), and male physiology-associated vulnerability (21).

### 4.4. Distinct treatment patterns of CVD

Despite a greater rate of hypertension, blood pressure-lowering medication was prescribed with slightly lesser intensity among females than males. This was previously reported by similar studies (22). In our study, the lower prescription concerned all hypertension drugs except for beta-blockers, which were taken more by females, perhaps due to other additional indications such as hyperthyroidism. According to data from Western/European countries, females tend to receive more thiazide diuretics while males display higher prescription of ACE inhibitors/ARBs and calcium-channel blockers but also a more frequent treatment interruption (23,24). Our male participants had more elevated SBP independent of age and BMI which suggests worse control of blood pressure. Moreover, compared to males, females reported receiving less oral antidiabetic therapy but higher oral antidiabetics plus insulin combination and insulin alone therapy. These observations may suggest a more frequent insulin-dependent diabetes among females than males. This is consistent with the notion of lower HbA1c reductions are obtained in women than in men for the same insulin dose (25).

### 4.5. Strengths, limitations, and implications for interventions

This is the largest nationwide study to evaluate the sex-related differences in CVD among Algerian individuals. Data were collected by a multidisciplinary team after first-hand extensive cardiometabolic evaluation. The observed results are therefore driven by a real-world assessment of the cardiovascular outcomes, and thus may reflect the status of CVD in Algeria and, by extension, the North Africa region.

On the other hand, various limitations are to be acknowledged. Participants were individuals who agreed to undergo cardiovascular screening. This may introduce an important self-selection bias, as subjects concerned about their health or those presenting cardiovascular risk factors may be more likely to participate. As such, the prevalence estimates could be subject to overestimation given the potential greater inclusion of subjects with preexisting cardiometabolic comorbidity. Moreover, because participants voluntarily attended a nationwide screening campaign, the cohort cannot be considered fully representative of the general population in Algeria. Thus, the prevalence estimates should be interpreted in light of the study context which is an opportunistic assessment of the records of healthcare-seeking individuals, who may differ from the general population, and thereby, the observed findings cannot be projected beyond this context. Besides selection bias, because of missing data, most of the analyses are based on variables with limited denominators therefore, the findings remain descriptive and are not generalized to the full cohort. Additionally, possible residual confounding, lack of socioeconomic variables, and the inability to infer causality were all notable weaknesses of the present study. Hence, given the observational design, it is not possible to identify the exact etiologies of sex-related disparities and disentangle biological factors from social and healthcare-related influences.

This study’s results can guide the prevention and management strategies that target CVD in underrepresented populations, such as North Africans. Both females and males require a tailored approach to reduce cardiovascular risks and optimize the effectiveness of current guidelines for cardiovascular care. For instance, recent AHA guidelines have advocated for the need to promote female physical activity (11). Likewise, in the 2021 ESC Guidelines on cardiovascular disease prevention in clinical practice, special attention was given to the impact of sex on CVD, however more specific recommendations remain warranted (26). Parallel to this, future studies are needed to further clarify the influence of sex on cardiovascular health, both from clinical and biological aspects.

## 5. Conclusion

In a large cardiometabolic screening program, sex groups were not equally affected by CVD with females being more prone to metabolic and anthropometric anomalies specifically dyslipidemia and obesity alongside a higher risk for hypertension. On the other hand, males showed a greater prevalence of overall CVD, coronary and peripheral artery disease, and left ventricular dysfunction consisting with a worse ischemic heart disease profile. These observed features likely reflect multifaceted biological, cardiometabolic, sociodemographic, and healthcare differences across sex groups. Therefore, there is a need for more large-scale studies to assess the factors contributing to the sex gap in the prevention and management of CVD.

## Abbreviations

CVD: Cardiovascular Disease
SBP: Systolic Blood Pressure
DBP: Diastolic Blood Pressure
ECG: Electrocardiogram
aOR: Adjusted Odds Ratio
CI: Confidence Interval
BMI: Body Mass Index
IQR: Interquartile Range
AIC: Akaike Information Criterion
LV: Left Ventricle / Left Ventricular
HbA1c: Glycated Hemoglobin
LDL: Low Density Lipoprotein
HDL: High Density Lipoprotein
ARB: Angiotensin II Receptor Blocker
ACE: Angiotensin Converting Enzyme
MENA: Middle East and North Africa

## Author Contributions

Conceptualization, M.B. and Y.K.; methodology, M.B., R.B., Y.T., D.F., and Y.K.; software, M.B., R.B., Y.T. and D.F.; validation, D.F., H.B. and N.L.; formal analysis, M.B.; investigation, M.B., and Y.K.; resources, M.B.; data curation, R.B., Y.T. and D.F.; writing—original draft preparation, M.B. and Y.K; writing—review and editing, D.F., H.B., N.L. and Y.K.; visualization, M.B.; supervision, H.B., N.L., N.B. and Y.K.; project administration, M.B.; funding acquisition, M.B. All authors have read and agreed to the published version of the manuscript.

## Institutional Review Board Statement

The study was conducted in accordance with the Declaration of Helsinki and approved by the Ethics Committee of Algeria’s National Ministry of Health.

## Informed Consent Statement

Informed consent was obtained from all subjects involved in the study.

## Data Availability Statement

The data underlying this article can be shared on reasonable request to the corresponding author after permission from the National Ministry of Health.

## Acknowledgments

None.

---

**Supplementary Table 1:** Tested variable in multivariate analysis.

| **Tested variable in multivariate analysis** | **Female N** | **Male N** | **Total N** |
| --- | --- | --- | --- |
| CKD | 7,619 | 5,870 | **13,489** |
| Known hypertension | 8,072 | 5,958 | **14,030** |
| Known diabetes | 8,029 | 6,450 | **14,479** |
| Known dyslipidemia | 6,001 | 4,649 | **10,650** |
| Abdominal obesity | 4,662 | 3,889 | **8,551** |
| Smoking | 9,661 | 7,750 | **17,411** |
| Physical activity | 19 | 568 | **587** |
| Dyslipidemia risk factors | 9,529 | 7,284 | **16,813** |
| Known CV disease | 9,433 | 7,383 | **16,816** |
| Family history of hypertension | 6,742 | 5,327 | **12,069** |
| Family history of diabetes | 6,735 | 5,337 | **12,072** |
| Family history of dyslipidemia | 6,603 | 5,236 | **11,839** |
| Heart failure | 7,619 | 5,870 | **13,489** |
| Sitting SBP | 3,687 | 3,035 | **6,722** |
| LDL | 124 | 123 | **247** |
| Total cholesterol | 28 | 40 | **68** |
| Triglyceride | 81 | 40 | **121** |
| HDL | 53 | 70 | **123** |

**Supplementary Table 2:** The main covariates used in the multivariable models:

| **Covariate** | **Female N** | **Male N** | **Total N** |
| --- | --- | --- | --- |
| Age | 12,015 | 9,507 | **21,522** |
| Sex | 12,015 | 9,507 | **21,522** |
| BMI | 8,252 | 6,655 | **14,907** |
| Known diabetes | 8,029 | 6,450 | **14,479** |
| Known hypertension | 8,072 | 5,958 | **14,030** |
| Known dyslipidemia | 6,001 | 4,649 | **10,650** |
| Family history of hypertension | 6,742 | 5,327 | **12,069** |
| Family history of diabetes | 6,735 | 5,337 | **12,072** |
| Family history of dyslipidemia | 6,603 | 5,236 | **11,839** |

## References

1. Naghavi M, Kyu HH, A B, Aalipour MA, Aalruz H, Ababneh HS, et al. Global burden of 292 causes of death in 204 countries and territories and 660 subnational locations, 1990–2023: a systematic analysis for the Global Burden of Disease Study 2023. Lancet [Internet]. 2025 Oct 18;406(10513):1811–72. Available from: 10.1016/S0140-6736(25)01917-8

2. Jortveit J, Sandberg EL, Pripp AH, Halvorsen S. Time trends in adherence to guideline recommendations for anticoagulation therapy in patients with atrial fibrillation and myocardial infarction. Open Hear [Internet]. 2022 Apr 6;9(1):e001934. Available from: 10.1136/openhrt-2021-001934

3. Li J, Zhang J, Somers VK, Covassin N, Zhang L, Xu H. Trends and Disparities in Treatment and Control of Atherosclerotic Cardiovascular Disease in US Adults, 1999 to 2018. J Am Heart Assoc [Internet]. 2024 May 7;13(9):e032527. Available from: 10.1161/JAHA.123.032527

4. Freedman AA, Colangelo LA, Ning H, Borrowman JD, Lewis CE, Schreiner PJ, et al. Sex Differences in Age of Onset of Premature Cardiovascular Disease and Subtypes: The Coronary Artery Risk Development in Young Adults Study. J Am Heart Assoc [Internet]. 2026 Feb 3;15(3):e044922. Available from: 10.1161/JAHA.125.044922

5. Kumar M, Hu JR, Ali S, Khlidj Y, Upreti P, Ati L, et al. Sex disparities in outcomes of transcatheter aortic valve implantation-a multi-year propensity-matched nationwide study. Int J Cardiol [Internet]. 2025 Jan 1;418. Available from: 10.1016/j.ijcard.2024.132619

6. Satish M, Walters RW, Wenzl FA, Safford M, Kini V. Sex Differences in Outcomes of Young Adults Hospitalized With First Myocardial Infarction From 2011 to 2022. J Am Heart Assoc [Internet]. 2026 Feb 27;0(0):e46517. Available from: 10.1161/JAHA.125.046517

7. G. RJ, E.B. RJ. Sex Differences in Cardiovascular Consequences of Hypertension, Obesity, and Diabetes. JACC [Internet]. 2022 Apr 19;79(15):1492–505. Available from: 10.1016/j.jacc.2022.02.010

8. Najman JM, Kisely S, Scott JG, Ushula TW, Williams GM, Clavarino AM, et al. Gender differences in cardiovascular disease risk: Adolescence to young adulthood. Nutr Metab Cardiovasc Dis [Internet]. 2024;34(1):98–106. Available from: https://www.sciencedirect.com/science/article/pii/S093947532300385X

9. Lv Y, Cao X, Yu K, Pu J, Tang Z, Wei N, et al. Gender differences in all-cause and cardiovascular mortality among US adults: from NHANES 2005–2018. Front Cardiovasc Med [Internet]. 2024;Volume 11. Available from: https://www.frontiersin.org/journals/cardiovascular-medicine/articles/10.3389/fcvm.2024.1283132

10. Lilian M, Martha G, Mzee N, Jasmit S, Bernard G, Felix B, et al. Sex Differences in Clinical Characteristics, Treatment, and Outcomes of Cardiovascular Disease. JACC Adv [Internet]. 2026 Jan 1;5(1):102466. Available from: 10.1016/j.jacadv.2025.102466

11. Jerome GJiE of PAP for OCH in AASSF the AHA, Boyer WR, Bustamante EE, Kariuki J, Lopez-Jimenez F, Paluch AE, et al. Increasing Equity of Physical Activity Promotion for Optimal Cardiovascular Health in Adults: A Scientific Statement From the American Heart Association. Circulation [Internet]. 2023 Jun 20;147(25):1951–62. Available from: 10.1161/CIR.0000000000001148

12. Chaabane S, Chaabna K, Abraham A, Mamtani R, Cheema S. Physical activity and sedentary behaviour in the Middle East and North Africa: An overview of systematic reviews and meta-analysis. Sci Rep. 2020 Jun;10(1):9363.

13. Asgari S, Moazzeni SS, Azizi F, Abdi H, Khalili D, Hakemi MS, et al. Sex-Specific Incidence Rates and Risk Factors for Hypertension During 13 Years of Follow-up : The Tehran Lipid and Glucose Study. Glob Heart. 2020;15(1):1–13.

14. Rezaianzadeh A, Johari MG, Baeradeh N, Seif M, Hosseini SV. Sex differences in hypertension incidence and risk factors: a population-based cohort study in Southern Iran. BMC Public Health [Internet]. 2024;24(1):3575. Available from: 10.1186/s12889-024-21082-8

15. States U, Ostchega Y, Ph D, Fryar CD, Nwankwo T, Nguyen DT. Hypertension Prevalence Among Adults Aged 18 and Over. Centers Dis Control Prev. 2020;(364):2017–8.

16. Patel N, Mittal N, Wilkinson MJ, Taub PR. Unique features of dyslipidemia in women across a lifetime and a tailored approach to management. Am J Prev Cardiol [Internet]. 2024;18:100666. Available from: https://www.sciencedirect.com/science/article/pii/S2666667724000345

17. Ballena-Caicedo J, Zuzunaga-Montoya FE, Loayza-Castro JA, Vásquez-Romero LEM, Tapia-Limonchi R, De Carrillo CIG, et al. Global prevalence of dyslipidemias in the general adult population: a systematic review and meta-analysis. J Heal Popul Nutr [Internet]. 2025;44(1):308. Available from: 10.1186/s41043-025-01054-3

18. Lombardo M, Feraco A, Armani A, Camajani E, Gorini S, Strollo R, et al. Gender differences in body composition, dietary patterns, and physical activity: insights from a cross-sectional study. Front Nutr. 2024;11:1414217.

19. Kakibuchi A, Ito F, Takaoka O, Tahara N, Kawamata M, Yabumoto K, et al. Effects of 17β-estradiol and equilin on atherosclerosis development in female Apoeshl mice. Sci Rep [Internet]. 2025;15(1):24922. Available from: 10.1038/s41598-025-10494-0

20. Molina E, Clarence EM, Ahmady F, Chew GS, Charchar FJ. Coronary Artery Disease: Why We should Consider the Y Chromosome. Hear Lung Circ [Internet]. 2016;25(8):791–801. Available from: https://www.sciencedirect.com/science/article/pii/S1443950616000366

21. Corban MT, Prasad A, Gulati R, Lerman LO, Lerman A. Sex-specific differences in coronary blood flow and flow velocity reserve in symptomatic patients with non-obstructive disease. EuroIntervention. 2021 Jan;16(13):1079–84.

22. Ambrož M, Geelink M, Smits KPJ, de Vries ST, Denig P. Sex disparities in medication prescribing amongst patients with type 2 diabetes mellitus managed in primary care. Diabet Med. 2023 Jan;40(1):e14987.

23. Bager JE, Manhem K, Andersson T, Hjerpe P, Bengtsson-Boström K, Ljungman C, et al. Hypertension: sex-related differences in drug treatment, prevalence and blood pressure control in primary care. J Hum Hypertens [Internet]. 2023;37(8):662–70. Available from: 10.1038/s41371-023-00801-5

24. Ljungman C, Kahan T, Schiöler L, Hjerpe P, Hasselström J, Wettermark B, et al. Gender differences in antihypertensive drug treatment: results from the Swedish Primary Care Cardiovascular Database (SPCCD). J Am Soc Hypertens [Internet]. 2014;8(12):882–90. Available from: https://www.sciencedirect.com/science/article/pii/S1933171114007499

25. Gourdy P, Bonadonna RC, Freemantle N, Mauricio D, Müller-Wieland D, Bigot G, et al. Does Gender Influence the Effectiveness and Safety of Insulin Glargine 300 U/ml in Patients with Uncontrolled Type 2 Diabetes? Results from the REALI European Pooled Analysis. Diabetes Ther [Internet]. 2022;13(1):57–73. Available from: 10.1007/s13300-021-01179-8

26. Visseren FLJ, Mach F, Smulders YM, Carballo D, Koskinas KC, Bäck M, et al. 2021 ESC Guidelines on cardiovascular disease prevention in clinical practice. Eur Heart J. 2021 Sep;42(34):3227–337.

